# Clinical validation of large-scale functional assays: insights from 2,120 gene-truthset-assay evaluations

**DOI:** 10.64898/2026.07.10.26357766

**Authors:** Sophie Allen, Charlie F. Rowlands, Zeid Kuzbari, Alice Garrett, Miranda Durkie, George J. Burghel, Rachel Robinson, Alison Callaway, Joanne Field, Bethan Frugtniet, Sheila Palmer-Smith, Jonathan Grant, Judith Pagan, Elizabeth Johnston, Trudi McDevitt, Lowri Hughes, Laura Yarram-Smith, Peter Logan, Laura Reed, Katie Snape, Terri McVeigh, Helen Hanson, Frederick P. Roth, Lea M. Starita, Douglas M. Fowler, Rehan Villani, Amanda B. Spurdle, David J. Adams, Greg Findlay, Clare Turnbull, CanVIG-UK

## Abstract

**Background:** Clear guidance is lacking regarding how ‘truthset’ variants should be used for clinical validation of functional assays, namely determining the allocatable evidence points (EPs) towards clinical classification. It is argued that assays should be validated using truthsets of missense variants, as this is the variant type for which classification is most impacted by functional data. EPs will be influenced by both the number of available ‘truthset’ variants and their concordance with assay readouts.

**Methods:** We first reviewed 112 sets of ClinGen gene-specific classification specifications (CSPECs) to assess methodologies they applied for truthset assembly and clinical validation of assays.

We then proposed differing rules regarding variant type and stringency of classification by which truthsets might be assembled using ClinVar-extracted classifications. We then examined augmentation of ClinVar-classified truthsets with ‘proxy-clinical’ benign-classified missense variants systematically assembled applying ACMG/AMP rules (of differing stringencies). In total, these constituted 70 basic approaches to ClinVar-based truthset assembly, which we applied to *VHL, BRCA1, BRCA2* and *RAD51C*. We additionally analysed the impact on the size of the truthsets of changing the specified phenotypes against which ClinVar classification had been submitted. We then applied these truthsets to quantify concordance and allocatable EPs for five large-scale multiplexed functional assays for *VHL*, *BRCA1*, *BRCA2*, and *RAD51C*.

**Results:** The EPs from clinical validation of each assay varied widely according to which truthset was used across 2,120 permutations of gene-truthset-assay combinations. For example, sequentially applying 700 different ClinVar-based truthsets to 2,268 VHL assay variant readouts (70 basic ClinVar-based approaches, augmented by examining 5 different phenotypes for each basic approach, and separate validation against two defined deleterious zones), the evidence strength allocatable for pathogenicity ranged from nil to strong evidence (0.0 to 5.6 EPs); for benignity it ranged from supporting to strong evidence (-1.5 to -6.4 EPs). Clinical validation using truthsets comprising just ClinVar-classified missense variants typically resulted in lower EPs than truthsets comprising protein truncating (PTV) and synonymous variants; this was more due to paucity of ClinVar-classified missense truthset variants than poorer concordance. Augmentation with larger ‘proxy-clinical’ benign-classified missense truthsets typically improved evidence allocatable for pathogenicity, with improved power negating modest reduction in concordance.

**Conclusions:** EPs can be improved by augmentation with systematically-generated ‘proxy-clinical’ benign-classified missense variants and/or reduction of truthset stringency. Explicit prescriptive clinical guidance is urgently required to improve consistency in clinical validation of functional assays and consequent evidence application for clinical variant classification.

## Background

A functional assay is an experiment that measures how a specific genetic variant affects a gene’s biological activity or product. Functional data from assays can be calibrated, with deleterious (non-functional, abnormal) readouts providing evidence of variant pathogenicity and neutral (functional, normal) readouts providing evidence of benignity in assays of tumour suppressor genes. In their 2015 variant classification framework (now termed v3), the American College of Medical Genetics and Genomics (ACMG)/Association for Molecular Pathology (AMP) articulated recommendations regarding assessment of how closely the assay reflected the in vivo environment, and robustness and validation of the data outputs, surmising that, a “well-established, robust, validated assay” was considered suitable for ‘strong’ evidence in variant classification(1). In 2020, in a publication by Brnich et al, the v3 guidance was extended by the ClinGen Sequence Variant Interpretation group, which produced a framework to quantify the evidence strength allocatable to an assay based on the concordance of assay results against a ‘truthset’ of previously classified variants (also termed in the literature as a ‘control-set’ or ‘clinical reference-set’)(2). In this ClinGen assay-level clinical validation framework, user-supplied threshold(s) (typically those provided by the assayist) are used to define functional classes (typically deleterious and neutral, +/- an intermediate zone). Then by assessing the functional classes assigned to the reference variants, numeric assay-level evidence strengths are generated for each functional class (termed Odds-Path, effectively representing Likelihood Ratios). Taking the log_2.08_ of these likelihood ratios generates allocatable ACMG/AMP evidence points (hereafter EPs) for pathogenicity and benignity(3, 4). These EPs are then applicable to all other variants with assay readouts above or below the user-supplied threshold(s).

With regard to assembly of these ‘truthset’ (control, reference) variants, Brnich et al state broadly that “controls should also be relevant to the disease mechanism (such as gain-of-function or loss-of-function) and the type of variant under consideration (e.g., missense controls for evaluating missense variants of uncertain significance)”(5). Increasingly large volumes of functional data from multiplex assays of variant effects (MAVEs) are being generated, with potential to have a substantial impact on variant classification (6). However, lack of clarity and consistency in the standards for clinical validation truthsets have been expressed as major hurdles for clinical diagnostic laboratories in using these MAVE data (7–9).

We thus sought to explore how different approaches to truthset assembly might impact the EPs assignable to functional assays. We first reviewed the Criteria Specification (CSPEC) Registry for 112 gene guidelines, to examine the approach adopted by 39 VCEPs for truthset assembly applied for clinical validation of functional assays. We then sought to examine factors influencing the size of truthset that could be assembled exclusively from ClinVar classifications, by applying differing stipulations regarding variant type, stringency of classification, and phenotype against which a variant had been submitted. We next evaluated augmentation of ClinVar-classified variants with ‘proxy-clinical’ benign-classified missense variants assembled by systematically applying ACMG/AMP v3.0 rules. We examined the resultant size of truthsets generated from these approaches for four cancer susceptibility genes (*VHL, BRCA1, BRCA2* and *RAD51C)*, all with loss-of-function mechanism of pathogenicity. We then quantified the EPs from application of these truthsets for clinical validation of corresponding large-scale functional assays for the four genes (10–14).

## Methods

### Criteria Specification (**CSPEC**) Registry Review

CSPEC pages and associated publications were reviewed for 112 genes across 39 VCEPs on 15^th^ December 2025. For each set of guidelines, PS3 and BS3 codes were reviewed for a) if the VCEP recommend application of the functional evidence codes, b) if any assays have been recommended by the VCEP, and c) if the VCEP have provided information on the variant set(s) used to validate the assays.

### Functional Assay Data sources

Function scores were sourced for 2,268 *VHL*, 3,893 *BRCA1*, 7,302 *BRCA2* (6,551 from Sahu et al., 2025, 6,959 from Huang et al., 2025) and 10,694 *RAD51C* variants assayed as previously published (10–14). Function scores from Huang et al., 2025 had been adjusted following processing by the VarCall pipeline as described, which was trained on all protein truncating (PTV)/synonymous variants with a SpliceAI score <0.2 (14).

All variants were annotated with the following information: variant information from Ensembl Variant Effect Predictor (VEP)(15) (v111) using the respective MANE Select transcript (*VHL*: ENST00000256474, *BRCA1*: ENST00000357654, *BRCA2*: ENST00000380152, *RAD51C*: ENST00000337432), population allele frequency from gnomAD v4.1(16), and predictive scores from REVEL(17), BayesDel noAF(18), and SpliceAI(19).

### Systematically-classified variant ‘truthsets’

We systematically assembled and classified protein truncating variants, synonymous, and missense variants to generate pathogenic and benign truthsets. Pathogenic truncating variants were defined as variants with an Ensembl VEP consequence including “stop_gained”, “splice_acceptor_variant”, “splice_donor_variant”, “frameshift_variant”, or “start_lost”, with a REVEL score ≥0.644 or BayesDel noAF score ≥0.13(20), and met the criteria for both PM2 and PVS1 per the respective variant curation expert groups (ClinGen *VHL* VCEP v1.1.0 (https://cspec.genome.network/cspec/ui/svi/affiliation/50036, ClinGen *BRCA1* and *BRCA2* VCEP v1.2.0 (https://cspec.genome.network/cspec/ui/svi/affiliation/50087), draft ClinGen *RAD51C* VCEP https://cspec.genome.network/cspec/ui/svi/affiliation/50039)). Benign synonymous variants were defined as variants with a “synonymous_variant” VEP consequence and a maximum Splice AI score ≤ 0.1.

Missense variants were defined as variants with a “missense_variant” VEP consequence. Benign missense variants were further defined using the methodology and inclusion criteria presented previously(21), using the thresholds as defined by the respective gene VCEP, and the allele frequency threshold for BS1_sup defined as half of the allele frequency threshold for BS1. The following rules identified four sets of benign missense truthset variants (with a minimum requirement of 2 observations in gnomAD v4.1 for a variant to be included in any truthset):

- Level 1: Attains BA1 (*VHL*: AF≥0.0156%; *BRCA1*: AF≥0.0868%; *BRCA2*: AF≥0.0906%; *RAD51C*: AF≥0.0583%) in any non-founder population in gnomAD v4.1
- Level 2: Attains BS1_strong (*VHL*: AF≥0.00156%; *BRCA1*: AF≥0.00868%; *BRCA2*: AF≥0.00906%; *RAD51C*: AF≥0.00583%) in any non-founder population in gnomAD v4.1 and benign evidence from either REVEL (≤0.290) or BayesDel noAF (≤0.15 for *BRCA1* and ≤0.18 for *BRCA2* following the ClinGen VCEP-calibrated thresholds for BP4, and ≤-0.18 for the remaining assays using the Pejaver thresholds for benign evidence(20)).
- Level 3: Attains BS1_sup (*VHL*: AF≥0.00078%; *BRCA1*: AF≥0.00434%; *BRCA2*: AF≥0.00453%; *RAD51C*: AF≥0.00292%)) in any non-founder population in gnomAD v4.1 **<u>and</u>** benign evidence from at least one predictive tool.
- Level 4: Attains BS1_sup (*VHL*: AF≥0.00078%; *BRCA1*: AF≥0.00434%; *BRCA2*: AF≥0.00453%; *RAD51C*: AF≥0.00292%) in any non-founder population in gnomAD v4.1 **<u>or</u>** benign evidence from at least one predictive tool.

### ClinVar-classified variant ‘truthsets’

ClinVar variant information files were downloaded to contain the most recent data archive available prior to assay publication date (*VHL*: 30/06/2024; *BRCA1*: 05/08/2018; *BRCA2*: 06/01/2025 for both assays; *RAD51C*: 01/10/2024)(22). 941 *VHL*, 913 *BRCA1*, 2,751 *BRCA2* (Sahu et al), 2,872 *BRCA2* (Huang et al), and 2,118 *RAD51C* variants were identified with both a ClinVar classification and corresponding assay function (Table S1). Analyses were also repeated for all genes using ClinVar variant data downloaded on 2^nd^ June 2025(22) (Table S1,S2).

### ClinGen assay-level clinical validation

Using the assayist-defined thresholds for deleterious (or non-functional) and neutral (or functional), clinical validation was conducted based on the approach described by Brnich et al., 2020(2), where:

- Prior, P1 is the proportion of pathogenic variants in the combined pathogenic and benign truthsets
- Posterior, P2:
- `o` For deleterious readouts: True Positives / (All deleterious readouts + 1)
- `o` For neutral readouts: (False Negatives +1) / (All neutral readouts + 1)

The Likelihood Ratio (“OddsPath”) towards pathogenicity (PS3) and benignity (BS3) was then calculated as per Tavtigian et al(3), where *P*_1_ is the prior probability and *P*_2_ is the posterior probability:

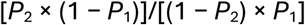

Likelihood Ratios (OddsPath) were then categorised according to ACMG/AMP v3 evidence strength categories by log transforming to base 2.08(4) to create a Log Likelihood Ratio, of EPs (where for evidence in either direction: supporting 1 EP, moderate 2 EPs, strong 4 EPs, and very strong 8 EPs).

We also calculated metrics to quantify concordance between truthsets and assay readouts, predicated on neither (i) truthset size (power) nor (ii) the relative proportions of pathogenic and benign truthset variants. The FalsePositiveRate_pathogenic_ is the number of neutral pathogenic-classified variants / total pathogenic-classified variants (akin to 1-Sensitivity). The FalsePositiveRate_benign_ is the number of deleterious benign-classified variants / total number benign-classified variants (akin to 1-Specificity). The variants scoring as functionally intermediate (ie not fully misclassified by the assay) were excluded from the denominators.

## Results

### Truthset construction is inconsistent across VCEPs

To assess the consistency in how ClinGen’s Variant Curation Expert Panels were generating and applying truthsets for clinical validation we reviewed 112 ClinGen gene-specific classification specifications (CSPECs). For 105 of 112, application of functional evidence was allowed with allowance of both PS3 and BS3 in 41% (43/105) and of PS3 only in 59% (62/105). Where functional evidence was allowed, 75% (79/105) recommended specific functional assays, of which the truthsets used in clinical validation were elucidated for only 61% (48/79) of guidelines (Table S3). Of these, information regarding the provenance of truthset variants was only provided for 25/48 (52%); 11/48 (23%) used a set of variants curated by the VCEP, a further 2/48 (4%) specified that the variants were previously classified by members of the VCEP via multifactorial-analyses, 1/48 (2%) used variants identified in cases, 2/48 (4%) used ClinVar-classified variant only, 1/48 (2%) used other variant databases and 8/48 (17%) used the variants included in the assay publication. Information about variant type and stringency of classification for the truthsets was available for only 2/48. This survey of CSPEC practice highlights 1) inconsistency and lack of transparency across expert panels regarding truthset assembly, 2) the consequent value of in-depth analysis into how truthset assembly affects the evidence strength that can be assigned to functional data and 3) the need for clearer guidance.

### Truthsets for a single gene can be constructed in a multitude of ways

We examined next the size of truthsets assembled from ClinVar-classified variants by applying different rules. By combining enumerations of variant type (all variants, versus only PTV and synonymous truthsets, versus missense-only), classification stringency including class (P and B only versus P/LP and B/LB) and number of stars (any versus ≥1 versus ≥2 versus ≥3), we recognised 24 approaches by which truthsets could be assembled. Furthermore, 0 and 1 LP and LB ClinVar classifications may include ‘soft-conflicts’ (e.g. where one submitter assigned as ‘VUS’ and one submitter assigned as ‘LP’); allowing or disallowing such variants increased the number of possible permutations to 30 approaches (Figure 1).

**Figure 1:**
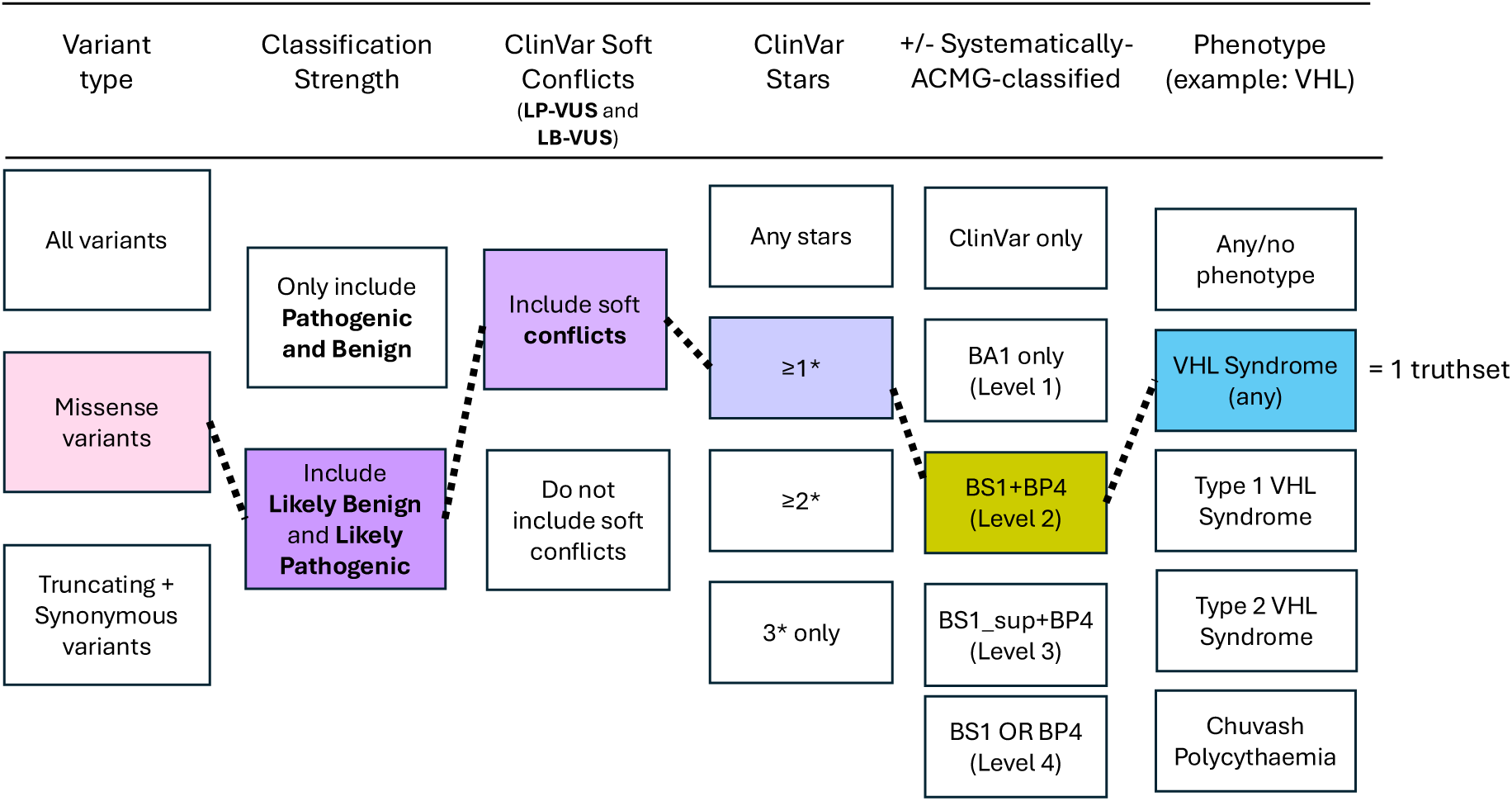
Permutations for assembly of ClinVar-based truthsets.

Next, for the 10/30 ‘missense-only’ approaches, we examined augmentation of ClinVar-classified benign missense variants with ‘proxy-clinical’ benign-classified missense variants systematically assembled by applying ACMG/AMP rules, considering four levels of stringency for combining evidence on variant frequency (BA1/BS1) and in silico predictions (BP4). This increased the number of permutations to 70 ClinVar-based approaches for truthset assembly; the size of pathogenicity and benignity truthsets generated is illustrated in Tables 1-3.

**Table 1:** Comparison of systematically-ACMG-classified PTV-synonymous truthsets against ClinVar-classified truthsets of different variant types. PTV Protein Truncating Variant; Syn Synonymous variant; P/LP Pathogenic or Likely Pathogenic; B/LB Benign or Likely Benign; EP ACMG Evidence Points. (1) Systematically-ACMG-classified: Truncating variant which is eligible for PVS1 and PM2 per VCEP specifications, and attains predictive evidence towards pathogenicity (using REVEL or BayesDel, noAF). (2) Systematically-ACMG-classified: Synonymous variant per VEP protein impact, and maximum SpliceAI score is ≤0.1

| Gene | Variant Type | Source (pathogenic) | Source (benign) | ClinVar class | ClinVar Soft-Conflicts | ClinVar stars | Truthset variants |  | FalsePositiveRate<sub>pathogenic</sub> | FalsePositiveRate<sub>benign</sub> | Balanced accuracy | Deleterious, Pathogenic | Deleterious, Benign | Intermediate, Pathogenic | Intermediate, Benign | Neutral, Pathogenic | Neutral, Benign | EP_PS3 | EP_BS3 |
| --- | --- | --- | --- | --- | --- | --- | --- | --- | --- | --- | --- | --- | --- | --- | --- | --- | --- | --- | --- |
|  |  |  |  |  |  |  | Pathogenic | Benign |  |  |  |  |  |  |  |  |  |  |  |
| VHL<br>(Buckley et al) | PTVs + Syn | Sys-ACMG-classified(1) | Sys-ACMG-classified(2) | NA | NA | NA | 68 | 369 | 0.03 | 0.01 | 0.98 | 66 | 5 | 0 | 10 | 2 | 354 | 5.58 | -4.20 |
|  | PTVs + Syn | ClinVar | ClinVar | P/LP and B/LB | Yes | ≥1 | 48 | 189 | 0.04 | 0.03 | 0.97 | 46 | 5 | 0 | 5 | 2 | 179 | 4.65 | -3.71 |
|  | Missense | ClinVar | ClinVar | P/LP and B/LB | Yes | ≥1 | 181 | 25 | 0.22 | 0.04 | 0.87 | 133 | 1 | 11 | 1 | 37 | 23 | 3.03 | -2.02 |
|  | All | ClinVar | ClinVar | P/LP and B/LB | Yes | ≥1 | 241 | 270 | 0.18 | 0.03 | 0.89 | 186 | 8 | 13 | 13 | 42 | 249 | 4.29 | -2.24 |
| BRCA1<br>(Findlay et al) | PTVs + Syn | Sys-ACMG-classified(1) | Sys-ACMG-classified(2) | NA | NA | NA | 45 | 504 | 0.00 | 0.00 | 1.00 | 45 | 2 | 0 | 8 | 0 | 494 | 7.00 | -5.17 |
|  | PTVs + Syn | ClinVar | ClinVar | P/LP and B/LB | Yes | ≥1 | 166 | 130 | 0.03 | 0.01 | 0.98 | 156 | 1 | 5 | 5 | 5 | 124 | 5.62 | -4.47 |
|  | Missense | ClinVar | ClinVar | P/LP and B/LB | Yes | ≥1 | 90 | 40 | 0.04 | 0.03 | 0.97 | 81 | 1 | 6 | 0 | 3 | 39 | 3.95 | -4.22 |
|  | All | ClinVar | ClinVar | P/LP and B/LB | Yes | ≥1 | 287 | 257 | 0.04 | 0.01 | 0.98 | 265 | 3 | 12 | 6 | 10 | 248 | 5.58 | -4.40 |
| BRCA2<br>(Sahu et al) | PTVs + Syn | Sys-ACMG-classified(1) | Sys-ACMG-classified(2) | NA | NA | NA | 228 | 1251 | 0.04 | 0.07 | 0.94 | 213 | 83 | 5 | 45 | 10 | 1123 | 3.59 | -3.99 |
|  | PTVs + Syn | ClinVar | ClinVar | P/LP and B/LB | Yes | ≥1 | 279 | 559 | 0.08 | 0.06 | 0.93 | 246 | 31 | 11 | 23 | 22 | 505 | 3.73 | -3.27 |
|  | Missense | ClinVar | ClinVar | P/LP and B/LB | Yes | ≥1 | 121 | 321 | 0.09 | 0.07 | 0.92 | 106 | 20 | 5 | 16 | 10 | 285 | 3.54 | -3.11 |
|  | All | ClinVar | ClinVar | P/LP and B/LB | Yes | ≥1 | 429 | 904 | 0.08 | 0.06 | 0.93 | 378 | 54 | 17 | 39 | 34 | 811 | 3.65 | -3.27 |
| BRCA2<br>(Huang et al) | PTVs + Syn | Sys-ACMG-classified(1) | Sys-ACMG-classified(2) | NA | NA | NA | 357 | 1265 | 0.02 | 0.00 | 0.99 | 349 | 2 | 2 | 1 | 6 | 1262 | 8.22 | -5.37 |
|  | PTVs + Syn | ClinVar | ClinVar | P/LP and B/LB | Yes | ≥1 | 304 | 571 | 0.02 | 0.00 | 0.99 | 296 | 2 | 2 | 3 | 6 | 566 | 7.13 | -5.14 |
|  | Missense | ClinVar | ClinVar | P/LP and B/LB | Yes | ≥1 | 116 | 310 | 0.04 | 0.05 | 0.96 | 109 | 16 | 3 | 3 | 4 | 291 | 3.88 | -4.21 |
|  | All | ClinVar | ClinVar | P/LP and B/LB | Yes | ≥1 | 453 | 994 | 0.02 | 0.02 | 0.98 | 437 | 20 | 5 | 13 | 11 | 961 | 5.22 | -4.91 |
| RAD51C<br>(Olvera-León et al) | PTVs + Syn | Sys-ACMG-classified(1) | Sys-ACMG-classified(2) | NA | NA | NA | 131 | 1599 | 0.07 | 0.02 | 0.95 | 122 | 38 | NA | NA | 9 | 1561 | 4.97 | -3.48 |
|  | PTVs + Syn | ClinVar | ClinVar | P/LP and B/LB | Yes | ≥1 | 213 | 527 | 0.02 | 0.02 | 0.98 | 209 | 9 | NA | NA | 4 | 518 | 5.39 | -5.10 |
|  | Missense | ClinVar | ClinVar | P/LP and B/LB | Yes | ≥1 | 29 | 108 | 0.14 | 0.06 | 0.90 | 25 | 6 | NA | NA | 4 | 102 | 3.53 | -2.32 |
|  | All | ClinVar | ClinVar | P/LP and B/LB | Yes | ≥1 | 261 | 835 | 0.03 | 0.03 | 0.97 | 253 | 25 | NA | NA | 8 | 810 | 4.69 | -4.56 |

*VHL* presented a use case for exploration of the impact of phenotype of submission as *VHL* deleterious variants are associated with several well-delineated phenotypes; in the heterozygous context Von Hippel Lindau cancer predisposition syndrome of various subtypes and in the biallelic context Chuvash polycythaemia. There existed in ClinVar 36 phenotypes (or ‘conditions’) against which classifications for *VHL* variants had been submitted (with additional permutations extractable from the free text, namely Type 1 and Type 2 Von Hippel Lindau Syndrome). Considering five phenotypes for variant submission, namely (i) any (or no) phenotype (ii) any ‘*VHL* syndrome’ phenotype and (iii) ‘Type 1 *VHL* syndrome’, (iv) ‘Type 2 *VHL* syndrome’, and (v) Chuvash Polycythaemia, the number of approaches for ClinVar-based truthset construction for *VHL* variants was scaled from 70 to 350 (Figure 1, Table 4).

### For clinical validation of an assay, different truthsets yield a range of allocatable EPs

We next used these assembled truthsets to validate published large-scale cellular fitness assays for *VHL, BRCA1, BRCA2* and *RAD51C*. For each clinical validation analysis, we calculated the allocatable EPs, as well as the FalsePositiveRate_benign_ (reflecting the assay specificity, the likelihood of a benign-classified truthset variant being assigned by the assay as deleterious) and the FalsePositiveRate_pathogenic_ (reflecting the assay sensitivity, the likelihood of a pathogenic-classified truthset variant being assigned by the assay as neutral).

We first bench-marked assay performance using ‘proxy-clinical’ benign-classified and pathogenic-classified truthsets comprising systematically-classified PTV plus synonymous variants (Table 1). Against these truthsets, each assay would attain allocatable evidence of ≥3 EPs towards both pathogenicity and benignity. The *BRCA2* Sahu et al. assay attained the weakest EPs_pathogenicity_ of 3.59 and concomitant highest FalsePositiveRate_benign_ (7%). The *RAD51C* assay attained the weakest EPs_benignity_ of -3.48, exhibiting the concomitant highest FalsePositiveRate_pathogenic_ (7%). Of note, the published scores from the Huang et al *BRCA2* assay had been pre-adjusted using a model trained on PTV-synonymous variants, thus inflating the apparent discriminatory performance for PTV versus synonymous truthsets.

We then performed clinical validation for each assay against each of the corresponding 70 ClinVar-based truthsets. The range of allocatable EPs (EPs) for each gene-assay dyad was broad (Tables 1-3, Table S1). For the Buckley et al *VHL* assay the EPs_pathogenicity_ ranged from -0.3 (rounded to 0.0, or nil evidence) to 5.6 and EPs_benignity_, -1.5 to -4.3 ; for the Findlay et al *BRCA1* assay EPs_pathogenicity_ were 1.9 to 6.0 and EPs_benignity_ -3.0 to -5.9, for the Sahu et al BRCA2 assay EPs_pathogenicity_ were 2.3 to 4.2 and EPs_benignity_ -1.4 to -4.1, for the Huang et al BRCA2 assay EPs_pathogenicity_ were 2.4 to 7.1 and EPs_benignity_ -2.9 to -7.1 and for the Olvera Leon et al RAD51C assay EPs_pathogenicity_ were 0.9 to 5.4 and EPs_benignity_ -2.2 to -6.8. This illustrated how for each assay using different truthsets could substantially alter the EPs allocatable.

### Applying missense-only truthsets typically results in lower allocatable EPs compared to PTV-synonymous truthsets

We examined three ‘variant type’ groupings of ClinVar-classified variants (i) just PTV and synonymous variants, (ii) just missense variants, or (iii) all available variants (Table 1). Considering ClinVarClinVar-classified truthsets of classifications of ≥1 star P/LP or B/LB including ‘all phenotypes’, across each of the five gene-assay dyads, the spread of assay scores for variants within missense truthsets were generally wider than for corresponding PTV pathogenicity truthsets or synonymous benignity truthsets; nevertheless the medians were generally comparable upon visual inspection (Figure 3, Table S4). Concordance measures (FalsePositiveRate_pathogenic_ and FalsePositiveRate_benign_) were variably modestly lower for ClinVar-classified missense-only compared to ClinVar-classified PTV-synonymous truthsets (with ‘all’ variants being intermediate). Notably, driven primarily by the relative paucity of available ClinVar-classified benign missense truthset variants, the EPs allocatable for missense-only clinical validation were typically substantially lower than for PTV-synonymous.

**Figure 3:**
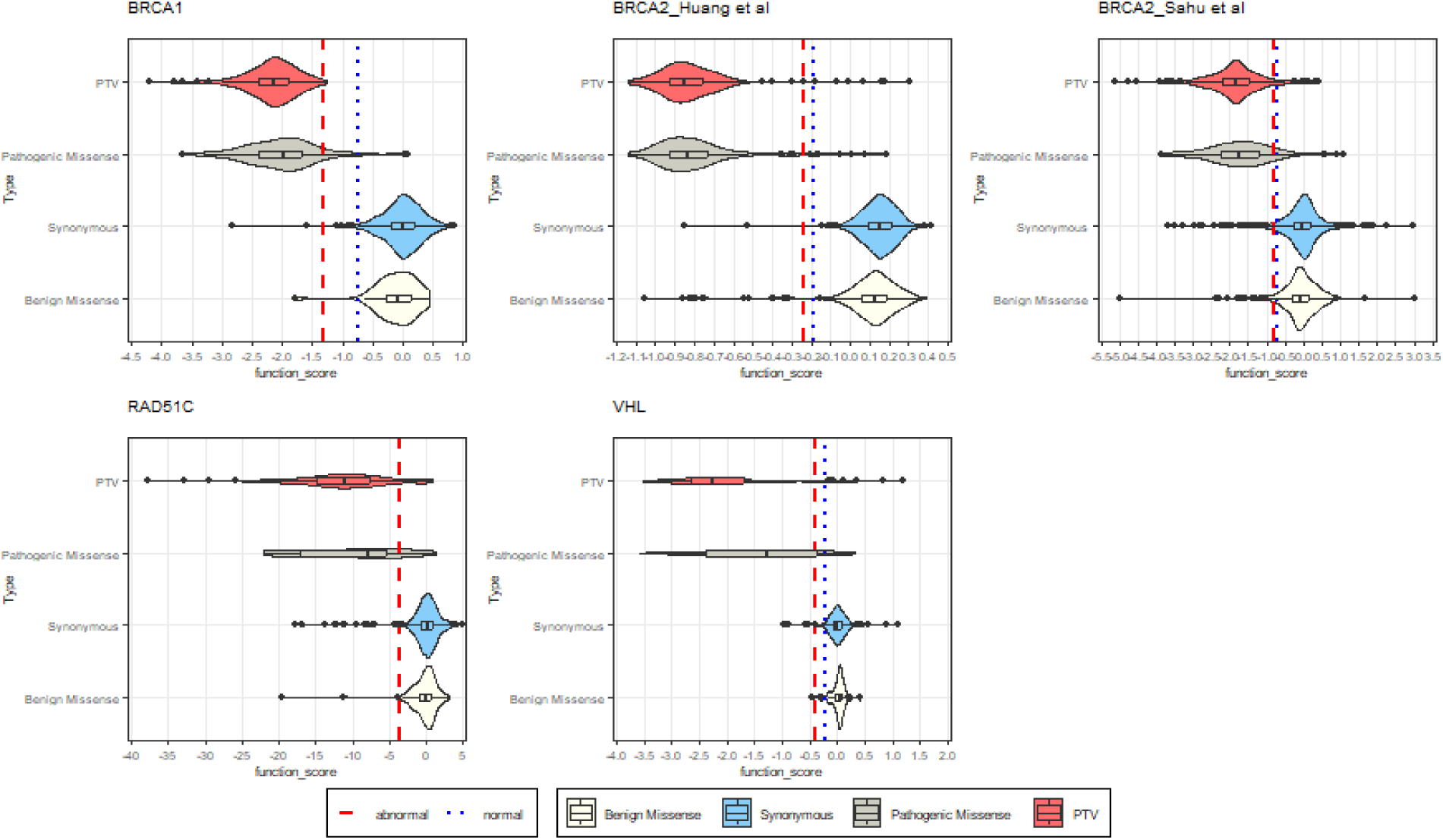
Comparison of score distributions for PTV and synonymous variants versus pathogenic and benign missense truthsets. Each missense truthset is sourced from ClinVar, are classified with ≥1 star, and include soft-conflicts. For *BRCA1, RAD51C*, and *VHL*, plotted thresholds for deleterious and neutral score ranges are sourced from the assay publication. For both *BRCA2* assays, as there are no discrete deleterious vs neutral thresholds, score thresholds are instead derived from the a) highest-scoring deleterious variant (for the ‘abnormal’ threshold), and b) lowest-scoring neutral variant (for the ‘normal’ threshold).

### Increasing the stringency of ClinVar-classifications typically reduces truthset size and EPs

We explored three metrics of classification stringency applicable to ClinVar extraction: classification strength (i.e. likely pathogenic vs pathogenic), number of submitters (i.e. number of stars) and allowance/non-allowance of soft-conflicts in underlying submitted classifications (i.e. variant classified as VUS by one submitter and LB by another) (Table 2, Table S1).

**Table 2:**
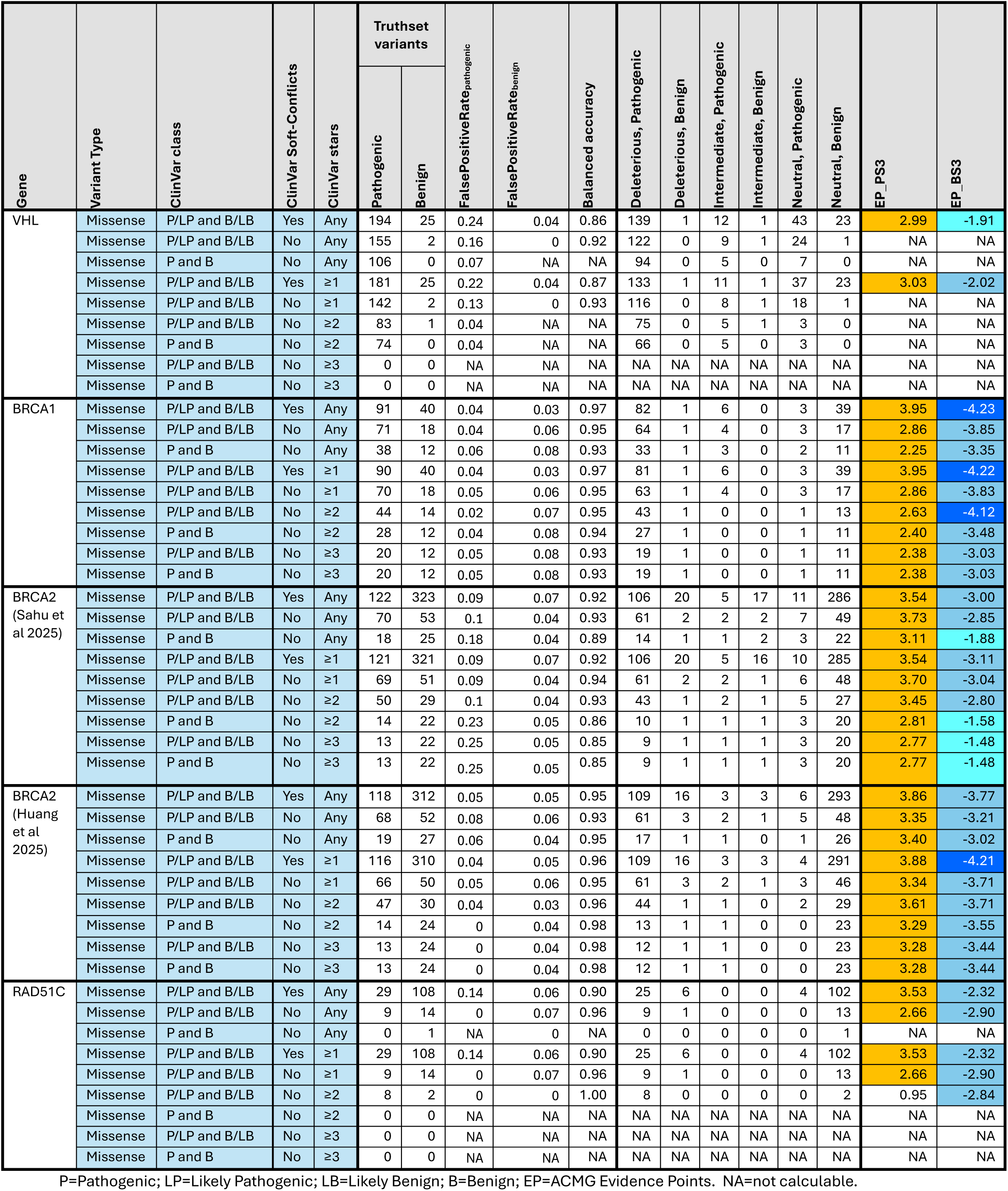
Comparison of ClinVar-classified truthsets comprising missense variants only.

For all assays examined, allowance of soft-conflicts dramatically increased the availability of truthset variants, in particular benign missense variants. Examining the impact of inclusion versus exclusion of soft-conflicts on concordance (rates of FalsePositiveRate_benign_ versus FalsePositiveRate_pathogenic)_ would imply that soft-conflicts of LP/VUS are more likely contaminated with true benign variants than LB/VUS are contaminated with true pathogenic variants. Thus, because of the paucity and because there is only a minor impact on FalsePositiveRate_benign_, inclusion of soft conflicts particularly enhances the EPs_pathogenicity_ from missense-only truthsets.

Greater stringency regarding (i) P/B versus P+LP/B+LB or (ii) higher number of stars resulted in modest improvement in concordance (FalsePositiveRate_pathogenic_ and FalsePositiveRate_benign_) but resulted in dramatic reduction in the number of truthset variants. Thus increased stringency typically resulted in reduced EPs (Table 2).

### Augmentation of ClinVar-classified variants with ‘proxy-clinical’ benign missense truthsets can improve allocatable EPs

We examined augmentation of ClinVar-classified classifications with ‘proxy-clinical’ benign-classified missense truthsets comprising variants systematically-classified via ACMG/AMP v3.0 rules. We explored four levels of stringency by which benignity was defined, relating to both population allele frequency (BA1/BS1) and *in silico* predictions (BP4, see methods). As anticipated, applying more stringent thresholds for systematic benignity classification resulted in improved concordance, whilst more permissive thresholds improved the number of variants generated. The balance of these influences varied by gene-assay dyad examined, with differing stringencies resulting in differing impact on maximum EPs_pathogenicity_ (Table 3).

**Table 3:** Comparison of ClinVar-based missense truthsets supplemented by systematically-ACMG-classified benign missense variants.

| Gene | Assembly of systematically ACMG/AMP classified truthsets | ClinVar class | ClinVar Soft-Conflicts | ClinVar stars | Truthset variants |  | FalsePositiveRate<sub>pathogenic</sub> | FalsePositiveRate<sub>benign</sub> | Balanced accuracy | Deleterious, Pathogenic | Deleterious, Benign | Intermediate, Pathogenic | Intermediate, Benign | Neutral, Pathogenic | Neutral, Benign | EP_PS3 | EP_BS3 |
| --- | --- | --- | --- | --- | --- | --- | --- | --- | --- | --- | --- | --- | --- | --- | --- | --- | --- |
|  |  |  |  |  | Pathogenic | Benign |  |  |  |  |  |  |  |  |  |  |  |
| VHL | Systematically-ACMG-classified 1 | P/LP and B/LB | Yes | ≥1 | 181 | 27 | 0.218 | 0.04 | 0.87 | 133 | 1 | 11 | 2 | 37 | 24 | 3.13 | -1.97 |
|  | Systematically-ACMG-classified 2 | P/LP and B/LB | Yes | ≥1 | 181 | 35 | 0.218 | 0.031 | 0.88 | 133 | 1 | 11 | 3 | 37 | 31 | 3.49 | -1.97 |
|  | Systematically-ACMG-classified 3 | P/LP and B/LB | Yes | ≥1 | 181 | 35 | 0.218 | 0.031 | 0.88 | 133 | 1 | 11 | 3 | 37 | 31 | 3.49 | -1.97 |
|  | Systematically-ACMG-classified 4 | P/LP and B/LB | Yes | ≥1 | 181 | 206 | 0.218 | 0.037 | 0.87 | 133 | 7 | 11 | 15 | 37 | 184 | 4.01 | -1.98 |
| BRCA1 | Systematically-ACMG-classified 1 | P/LP and B/LB | Yes | ≥1 | 90 | 40 | 0.036 | 0.025 | 0.97 | 81 | 1 | 6 | 0 | 3 | 39 | 3.95 | -4.22 |
|  | Systematically-ACMG-classified 2 | P/LP and B/LB | Yes | ≥1 | 90 | 45 | 0.036 | 0.023 | 0.97 | 81 | 1 | 6 | 1 | 3 | 43 | 4.11 | -4.19 |
|  | Systematically-ACMG-classified 3 | P/LP and B/LB | Yes | ≥1 | 90 | 47 | 0.036 | 0.022 | 0.97 | 81 | 1 | 6 | 2 | 3 | 44 | 4.17 | -4.16 |
|  | Systematically-ACMG-classified 4 | P/LP and B/LB | Yes | ≥1 | 90 | 1115 | 0.036 | 0.027 | 0.97 | 81 | 28 | 6 | 59 | 3 | 1028 | 4.84 | -4.14 |
| BRCA2<br>(Sahu et al) | Systematically-ACMG-classified 1 | P/LP and B/LB | Yes | ≥1 | 121 | 321 | 0.086 | 0.066 | 0.92 | 106 | 20 | 5 | 16 | 10 | 285 | 3.54 | -3.11 |
|  | Systematically-ACMG-classified 2 | P/LP and B/LB | Yes | ≥1 | 121 | 326 | 0.086 | 0.071 | 0.92 | 106 | 22 | 5 | 17 | 10 | 287 | 3.44 | -3.10 |
|  | Systematically-ACMG-classified 3 | P/LP and B/LB | Yes | ≥1 | 121 | 347 | 0.086 | 0.076 | 0.92 | 106 | 25 | 5 | 19 | 10 | 303 | 3.36 | -3.09 |
|  | Systematically-ACMG-classified 4 | P/LP and B/LB | Yes | ≥1 | 121 | 3571 | 0.086 | 0.104 | 0.90 | 106 | 352 | 5 | 200 | 10 | 3019 | 2.98 | -3.04 |
| BRCA2<br>(Huang et al) | Systematically-ACMG-classified 1 | P/LP and B/LB | Yes | ≥1 | 116 | 310 | 0.04 | 0.05 | 0.96 | 109 | 16 | 3 | 3 | 4 | 291 | 3.88 | -4.21 |
|  | Systematically-ACMG-classified 2 | P/LP and B/LB | Yes | ≥1 | 116 | 315 | 0.04 | 0.06 | 0.95 | 109 | 18 | 3 | 3 | 4 | 294 | 3.74 | -4.20 |
|  | Systematically-ACMG-classified 3 | P/LP and B/LB | Yes | ≥1 | 116 | 332 | 0.04 | 0.05 | 0.95 | 109 | 18 | 3 | 3 | 4 | 311 | 3.82 | -4.20 |
|  | Systematically-ACMG-classified 4 | P/LP and B/LB | Yes | ≥1 | 116 | 3564 | 0.04 | 0.06 | 0.95 | 109 | 201 | 3 | 63 | 4 | 3300 | 3.83 | -4.19 |
| RAD51C | Systematically-ACMG-classified 1 | P/LP and B/LB | Yes | ≥1 | 29 | 109 | 0.138 | 0.055 | 0.90 | 25 | 6 | 0 | 0 | 4 | 103 | 3.55 | -2.32 |
|  | Systematically-ACMG-classified 2 | P/LP and B/LB | Yes | ≥1 | 29 | 126 | 0.138 | 0.056 | 0.90 | 25 | 7 | 0 | 0 | 4 | 119 | 3.56 | -2.32 |
|  | Systematically-ACMG-classified 3 | P/LP and B/LB | Yes | ≥1 | 29 | 146 | 0.138 | 0.075 | 0.89 | 25 | 11 | 0 | 0 | 4 | 135 | 3.21 | -2.29 |
|  | Systematically-ACMG-classified 4 | P/LP and B/LB | Yes | ≥1 | 29 | 1682 | 0.138 | 0.109 | 0.88 | 25 | 183 | 0 | 0 | 4 | 1499 | 2.82 | -2.24 |

P/LP=Pathogenic or Likely Pathogenic; B/LB=Benign or Likely Benign; EP=ACMG Evidence Points. (1) Attains BA1 in any non-founder population in gnomAD v4.1. (2) Attains BS1\_strong in any non-founder population in gnomAD v4.1 and benign evidence from either REVEL ( $\leq 0.290$ ) or BayesDel noAF ( $\leq 0.15$ for BRCA1 and $\leq 0.18$ for BRCA2, $\leq -0.18$ for VHL and RAD51C). (3) Attains BS1\_sup in any non-founder population in gnomAD v4.1 and benign evidence from at least one predictive tool. (4) Attains BS1\_sup in any non-founder population in gnomAD v4.1 and/or benign evidence from at least one predictive tool.

### Restriction of phenotype of ClinVar submission will limit truthset size with impact on allocatable EPs

When ‘truthsets’ were restricted to *VHL* variants for which ClinVar pathogenicity classifications were reported against specific phenotypes, use of the more specific phenotypes of Type 1 and Type 2 von Hippel-Lindau Syndrome improved concordance (reducing the FalsePositiveRate_pathogenic_). However, this analysis illustrates how restricting the phenotype of ClinVar-submitted classifications reduces truthset size, meaning that using a more specific phenotype definition overall tended to reduce EPs_benignity_ despite the increase in overall concordance (Table 4).

**Table 4:** Comparison of ClinVar-classified truthsets of all VH variants using different phenotype associations.

| Gene | Variant Type | ClinVar class | ClinVar Soft-Conflicts | ClinVar stars | ClinVar Phenotype | Truthset variants |  | FalsePositiveRate<sub>pathogenic</sub> | FalsePositiveRate<sub>benign</sub> | Balanced accuracy | Deleterious, Pathogenic | Deleterious, Benign | Intermediate, Pathogenic | Intermediate, Benign | Neutral, Pathogenic | Neutral, Benign | EP_PS3 | EP_BS3 |
| --- | --- | --- | --- | --- | --- | --- | --- | --- | --- | --- | --- | --- | --- | --- | --- | --- | --- | --- |
|  |  |  |  |  |  | Pathogenic | Benign |  |  |  |  |  |  |  |  |  |  |  |
| VHL | All | P/LP and B/LB | Yes | ≥1 | Any | 241 | 270 | 0.2 | 0 | 0.89 | 186 | 8 | 13 | 13 | 42 | 249 | 4.29 | -2.24 |
|  | All | P/LP and B/LB | Yes | ≥1 | von Hippel-Lindau Syndrome | 222 | 246 | 0.2 | 0 | 0.89 | 170 | 6 | 12 | 12 | 40 | 228 | 4.50 | -2.20 |
|  | All | P/LP and B/LB | Yes | ≥1 | Type 1 von Hippel-Lindau Syndrome | 7 | 246 | 0.1 | 0 | 0.92 | 6 | 6 | 0 | 12 | 1 | 228 | 4.65 | -1.61 |
|  | All | P/LP and B/LB | Yes | ≥1 | Type 2 von Hippel-Lindau Syndrome | 21 | 246 | 0.1 | 0 | 0.93 | 16 | 6 | 3 | 12 | 2 | 228 | 4.49 | -2.55 |
|  | All | P/LP and B/LB | Yes | ≥1 | Chuvash Polycythaemia | 177 | 242 | 0.2 | 0 | 0.87 | 128 | 5 | 11 | 12 | 38 | 225 | 4.61 | -1.97 |

P/LP=Pathogenic or Likely Pathogenic; B/LB=Benign or Likely Benign; EP=ACMG Evidence Points.

### Allocatable EPs towards pathogenicity can be increased by tightening the deleterious (non-functional) score range

For both the *VHL* and *RAD51C* assays, the assay-authors had delineated two subcategories within the deleterious score range. For *VHL* these zones were entitled LOF1 and LOF2; for *RAD51C* the zones were entitled ‘fast-depletion’ and ‘slow-depletion’. As anticipated, restricting to the more extreme scoring zone for deleterious improved concordance (FalsePositiveRate_benign_) and consequently EPs_pathogenicity_. This analysis illustrates that such restriction inevitably results in a lower proportion of variants assigned as deleterious with more variants assigned to an indeterminate zone (Table 5).

**Table 5:** Comparison of truthsets for VH and RAD51C, with the function score zone for variants considered deleterious (or non-functional) defined using different thresholds.

| Gene | Deleterious zone | Variant Type | Source (pathogenic) | Source (benign) | ClinVar class | ClinVar Soft-Conflicts | ClinVar stars | Truthset variants |  | FalsePositiveRate<sub>pathogenic</sub> | FalsePositiveRate<sub>benign</sub> | Balanced accuracy | Deleterious, Pathogenic | Deleterious, Benign | Intermediate, Pathogenic | Intermediate, Benign | Neutral, Pathogenic | Neutral, Benign | EP_PS3 | EP_BS3 |
| --- | --- | --- | --- | --- | --- | --- | --- | --- | --- | --- | --- | --- | --- | --- | --- | --- | --- | --- | --- | --- |
|  |  |  |  |  |  |  |  | Pathogenic | Benign |  |  |  |  |  |  |  |  |  |  |  |
| VHL | LOF1 + LOF2 | PTVs + Syn | Sys-ACMG-classified(1) | Sys-ACMG-classified(2) | NA | NA | NA | 68 | 369 | 0.03 | 0.01 | 0.98 | 66 | 5 | 0 | 10 | 2 | 354 | 5.58 | -4.20 |
|  |  | PTVs + Syn | ClinVar | ClinVar | P/LP and B/LB | Yes | ≥1 | 48 | 189 | 0.04 | 0.03 | 0.97 | 46 | 5 | 0 | 5 | 2 | 179 | 4.65 | -3.71 |
|  |  | Missense | ClinVar | ClinVar | P/LP and B/LB | Yes | ≥1 | 181 | 25 | 0.22 | 0.04 | 0.87 | 133 | 1 | 11 | 1 | 37 | 23 | 3.03 | -2.02 |
|  |  | All | ClinVar | ClinVar | P/LP and B/LB | Yes | ≥1 | 241 | 270 | 0.18 | 0.03 | 0.89 | 186 | 8 | 13 | 13 | 42 | 249 | 4.29 | -2.24 |
|  | LOF1 only | PTVs + Syn | Sys-ACMG-classified(1) | Sys-ACMG-classified(2) | NA | NA | NA | 68 | 369 | 0.03 | 0.00 | 0.98 | 63 | 0 | 3 | 15 | 2 | 354 | 7.97 | -4.20 |
|  |  | All | ClinVar | ClinVar | P/LP and B/LB | Yes | ≥1 | 241 | 270 | 0.23 | 0.00 | 0.88 | 140 | 0 | 59 | 21 | 42 | 249 | 6.90 | -2.24 |
|  |  | Missense | ClinVar | ClinVar | P/LP and B/LB | Yes | ≥1 | 181 | 25 | 0.29 | 0.00 | 0.86 | 92 | 0 | 52 | 2 | 37 | 23 | 3.47 | -2.02 |
|  |  | PTVs + Syn | ClinVar | ClinVar | P/LP and B/LB | Yes | ≥1 | 48 | 189 | 0.04 | 0.00 | 0.98 | 44 | 0 | 2 | 10 | 2 | 179 | 7.04 | -3.71 |
| RAD51C | Fast + Slow depleters | PTVs + Syn | Sys-ACMG-classified(1) | Sys-ACMG-classified(2) | NA | NA | NA | 131 | 1599 | 0.07 | 0.02 | 0.95 | 122 | 38 | NA | NA | 9 | 1561 | 4.97 | -3.48 |
|  |  | PTVs + Syn | ClinVar | ClinVar | P/LP and B/LB | Yes | ≥1 | 213 | 527 | 0.02 | 0.02 | 0.98 | 209 | 9 | NA | NA | 4 | 518 | 5.39 | -5.10 |
|  |  | Missense | ClinVar | ClinVar | P/LP and B/LB | Yes | ≥1 | 29 | 108 | 0.14 | 0.06 | 0.90 | 25 | 6 | NA | NA | 4 | 102 | 3.53 | -2.32 |
|  |  | All | ClinVar | ClinVar | P/LP and B/LB | Yes | ≥1 | 261 | 835 | 0.03 | 0.03 | 0.97 | 253 | 25 | NA | NA | 8 | 810 | 4.69 | -4.56 |
|  | Fast depleters only | PTVs + Syn | Sys-ACMG-classified(1) | Sys-ACMG-classified(2) | NA | NA | NA | 131 | 1599 | 0.07 | 0.01 | 0.96 | 120 | 22 | 2 | 16 | 9 | 1561 | 5.67 | -3.48 |
|  |  | All | ClinVar | ClinVar | P/LP and B/LB | Yes | ≥1 | 261 | 835 | 0.03 | 0.02 | 0.98 | 243 | 13 | 10 | 12 | 8 | 810 | 5.48 | -4.56 |
|  |  | Missense | ClinVar | ClinVar | P/LP and B/LB | Yes | ≥1 | 29 | 108 | 0.17 | 0.03 | 0.90 | 19 | 3 | 6 | 3 | 4 | 102 | 3.92 | -2.32 |
|  |  | PTVs + Syn | ClinVar | ClinVar | P/LP and B/LB | Yes | ≥1 | 213 | 527 | 0.02 | 0.01 | 0.99 | 207 | 3 | 2 | 6 | 4 | 518 | 6.63 | -5.10 |

### Censoring the date of ClinVar extraction to match the date of assay publication can reduce truthset size

For our primary analysis, we used a ClinVar extract contemporaneous to the date of assay publication; however, we repeated the analyses using ClinVar-extracted classifications at the time of analysis (01/06/2025, Table S2). As might be predicted, in particular for the *BRCA1* assay published in 2018, this resulted in a marked increase in the number of available missense variant classifications. Furthermore, a higher number of variants had attained a Pathogenic/Benign classification (rather than Likely Pathogenic/Likely Benign) and an increased proportion of variants had attained 2 and 3 classifications. There was also higher concordance between truthset classifications and assay outputs for missense variants using the more recent ClinVar extract (reduced FalsePositiveRate_benign_ and FalsePositiveRate_pathogenic_), as would be anticipated if assay data had contributed to these more recent classifications. This analysis illustrates the circularity that arises when functional datasets may be driving variant classifications, highlighting the need for caution in using historical ClinVar extractions.

## Discussion

We have demonstrated first the wide heterogeneity in current VCEP practice in assembly of truthsets and clinical validation of functional assays via review of CSPECs for 112 genes from ClinGen VCEPs. We have demonstrated second the wide variation in size of truthsets possible from different rules using ClinVar-classified variants as well as additional permutations possible from augmenting ClinVar-classified truthsets with ‘proxy-clinical’ benign-classified missense truthsets systematically-classified via ACMG-AMP v3.0: we identified 70 approaches with non-exhaustive enumerations of variables and before considering vagaries of clinical phenotypes. Third, examining 2,120 truthset-gene-assay combinations in total, we illustrate across five corresponding large-scale functional assays how using these different approaches to constructing truthsets can generate widely varying allocatable evidence points (EPs) on this ClinGen assay-level clinical validation. These observations substantiate calls for more explicit standards defining how truthsets are constructed and methodology deployed for clinical assay validation and provide rich substrate for considering the challenges and complexity therein. (23).

### Implications of the ClinGen assay-level clinical validation framework and its methodology

Inherent to consideration of the assembly and performance of truthsets is appreciation of the mechanics of the ClinGen assay-level validation framework described by Brnich et al. The first important feature to note is incorporation of a (+1) to the denominator for the posterior probability (Haldane correction), with the stated intent of curbing infinity results and attenuation of the likelihood ratio (odds-path)(2). This attenuation will be proportionately more punitive with smaller truthsets.

The second important feature is the paradoxical relationship whereby the LikelihoodRatio_pathogenicity_ (and thus EPs_pathogenicity_) is primarily influenced by the size of benign truthsets whilst the LikelihoodRatio_benignity_ (and thus EPs_benignity_) is primarily influenced by the size of pathogenic truthsets. It is a consequence of these two features, that when undertaking clinical validation using a small benignity truthset, the allocatable EPs for pathogenicity (EPs_pathogenic_) is typically low even when concordance (FalsePositiveRate_benign_) is high.

The third consideration of the ClinGen assay-level validation methodology is reliance on the assayist-defined thresholds for neutral and deleterious, which have typically been derived in calibration using PTV-synonymous variants. Particularly where an assay is highly discriminatory, clean separation of PTV and synonymous could mean that the ‘intermediate zone’ derived via a maximum likelihood approach is extremely narrow with ‘bald’ zones in the assay score range. It is thus informative to examine the distribution of missense truthset variants in relation to the assayist-defined thresholds to compare their similarity to that of corresponding PTV-synonymous variant truthsets.

Finally, it is critical to appreciate that pathogenicity and benignity are not ‘symmetrical’ in regard of the clinical validation of functional assays. (i) An assay may capture only a subset of the mechanisms by which a variant can have a deleterious impact on the protein; ‘pathogenicity’ can be ascribed if a variant is demonstrated as deleterious from a single mechanism whilst assignation of benignity requires exclusion of all mechanisms. (ii) Conversely, for a variant with limited additional information (a “cold” VUS), application of functional data to enable classification as (likely) pathogenic results in actionability whilst “cold-VUS” to benign has no clinical impact. (iii) Evidence requirements are different: in the forthcoming ACMG/AMP v4.0, a likely benign classification will now be attainable with only one evidence point (from a single source) whilst ‘likely pathogenic’ requires six.

### The influence of truthset stringency on allocatable EPs

Relaxation of the stringency of ClinVar-extracted missense truthsets or via systematic assembly (i.e. greater contamination with incorrect classifications) will attenuate the EPs from clinical validation due to regression towards the null. However, the benefit in power afforded by the larger number of truthset variants may on many occasions counterbalance this deflation. Notably, relaxing stringency will only ever deflate the allocatable EPs for the assay compared to using a hypothetical ‘perfect truthset’ and thus can be embraced where the improved power translates into net improvement in allocatable EPs. The same principle of trade-off of concordance (precision) for power applies to use of a broader versus narrower phenotypic definition (providing all phenotypes are associated with the same mutational mechanism of action).

### The implications of using missense truthsets versus PTV-synonymous

Functional data will be most likely to influence classifications out of the VUS category for missense (and other non-truncating) variants; hence the implications of clinical validation on this category of variants is under most scrutiny. Thus, whilst it is a valuable first step to demonstrate that the assay distinguishes highly deleterious (PTVs) from neutral (synonymous) variants, the critical question is “Can the assay distinguish pathogenic missense variants from benign missense variants?” A MAVE could perfectly separate PTVs from synonymous variants and still perform poorly on missense variants if: many pathogenic missense variants are only partially damaging, benign missense variants produce modest functional perturbations, and/or the assay captures one aspect of protein function but not others. Furthermore, for assays not recapitulating genomic context, splicing effects and nonsense mediated decay are not captured. These factors form the basis of the argument that missense variants should be used for clinical validation.

As our data illustrate, there is frequently a paucity of ClinVar-classified missense truthset variants in particular where high stringency has been applied; whilst typically most acute for benign missense truthset variants we have illustrated that this issue can be mitigated by systematic construction thereof. A widely used practice to mitigate extreme paucity of ClinVar-classified pathogenic missense variants in order to empower allocatable EPs is to supplement with truncating variants (as illustrated in our analyses for ‘all’ ClinVar-classified variants). However, using a truthset comprising an arbitrary mixture of missense/PTV/synonymous variants is potentially problematic as (failure of) assay concordance for missense variants may be diluted by assay concordance for PTV-synonymous. Notably, recalling (i) the paradoxical relationship within the ClinGen assay-level validation framework and (ii) clinical utility largely lies in classifying variants as pathogenic and (iii) only one evidence point will be required for classification of benignity, that a paucity of pathogenic missense truthset variants results in only modest EPs for benignity may not be unduly compromising in regard of clinical variant classification.

Where there are sufficient missense-only benignity and pathogenicity truthset variants to undertake clinical validation of an assay, it should be ensured that the distribution of assay scores in relation to thresholds is broadly comparable to those for PTV-synonymous variants, as the latter will typically have been used to calibrate the assay to generate thresholds.

## Conclusion

Based on surveying 112 VCEP CSPEC pages and examination of 2,120 different truthset-gene dyads, we present a detailed exposition of the different approaches available for clinical validation. In particular, these analyses highlight the consequences and implications on allocatable EPs when ClinVar-extracted truthsets are restricted according to variant type, stringency and phenotype. These analyses also demonstrate how systematically-generated ‘proxy-clinical’ classifications of differing stringency can augment truthsets of benign missense variants, empowering the evidence allocatable for pathogenicity which is arguably of most clinical utility. Insights from these data will facilitate development of new, more prescriptive guidelines on assembly of truthset variants for clinical validation of functional assays.

## Availability of data and materials

The ClinVar datasets used and/or analysed during the current study are available in the ClinVar repository, https://ftp.ncbi.nlm.nih.gov/pub/ClinVar/. The functional assay datasets analysed during the current study are available in their respective published articles [and their supplementary information files] (*VH*L: https://doi.org/10.1038/s41588-024-01800-z(11)*; BRCA1* https://doi.org/10.1038/s41586-018-0461-z(10) and interim short report available at https://www.cangene-canvaruk.org/_files/ugd/ed948a_0399a952a1dc4767bed4364a04f6408b.pdf*; BRCA2* Sahu et al., 2025: https://doi.org/10.1038/s41586-024-08349-1(13); *BRCA2* Huang et al., 2025: https://doi.org/10.1038/s41586-024-08388-8(14);

*RAD51C:* https://doi.org/10.1016/j.cell.2024.08.039(12)). All other data generated or analysed during the current study are included in this published article [and its supplementary information files].

## Author contributions

Conceptualisation: CT, AS, FPR, LS, DA, GF; Data Curation: SA; Software: CFR, SA; Formal Analysis: SA, CFR; Funding acquisition: CT; Investigation: SA; Methodology: CT, SA, CFR, AG; Project Administration: SA, Supervision: CT; Visualisation: SA; Validation: MD, GJB, AC, RR, JF, BF, SPS, JG, JP, EJ, TMcD, LH, LY-S, PL, LR, KS, HH, TMcV, CanVIG-UK; Writing – original draft: CT, SA; Writing - review and editing: all authors.

## Acknowledgements

[TBC]

